# Systematic review of health state utility values used in pharmacoeconomic evaluations for chronic hepatitis C: impact on cost-effectiveness results

**DOI:** 10.1101/2020.06.22.20135434

**Authors:** Ru Han, Clément François, Mondher Toumi

## Abstract

**Background:** Health state utility values (HSUVs) identified from utility elicitation studies are widely used in pharmacoeconomic evaluations for chronic hepatitis C (CHC) and are particularly instrumental in health technology assessment (HTA) evaluation like the National Institute for Health and Clinical Excellence (NICE).

**Objective:** The objective of this study is to identify HSUVs used in cost-utility analyses (CUAs) for CHC in Europe and evaluate the impact of HSUVs selection on cost-effectiveness results in terms of incremental cost per quality-adjusted life-year (QALY) gained (ICER).

**Methods:** A systematic search of pharmacoeconomic evaluations for CHC was updated in Medline and Embase from the period of 2012-2017 to the period of 2017-2020. Data on health states, HSUVs and utility elicitation studies were extracted. The difference in HSUVs of the same health state in different CUAs and the difference between HSUVs of one health state and of the interlink health state in the same CUAs were calculated. A quality assessment was performed to evaluate the selection of HSUVs in CUAs. Sets of HSUVs identified were used in a re-constructed CUA model to assess the impact on ICER.

**Results:** Twenty-six CUAs conducted in European countries and referring to 17 utility elicitation studies were included. The difference in HSUVs of the same health states in different CUAs ranged from 0.021 (liver transplant) to 0.468 (decompensated cirrhosis). The difference between HSUVs of one health state and of the interlink health state of next disease severity level was calculated between health state of F0-F1/mild and F2-F3/moderate (n=11, 0.040 to 0.110), F2-F3/moderate and F4/compensated cirrhosis (n=18, 0.027 to 0.130), compensated cirrhosis and decompensated cirrhosis (n=22, 0.020 to 0.100), decompensated cirrhosis and hepatocellular carcinoma (n=24, 0.000 to 0.200), hepatocellular carcinoma and liver transplant in the first year (n=17, −0.329 to 0.170) and liver transplant in the first year and in subsequent years (n=17, −0.340 to 0.000). The utility elicitation study selected by most CUAs (n=11)was recommended as the source of HSUVs, as least for the CUAs conducted in the UK, based on the results of quality assessment. Seven sets of HSUVs were generated to fit the re-constructed model and changed the results of incremental analysis from being cost-effective to not cost-effective (ICER raging from £2,460 to £24,954 per QALY gained), and to dominated in the UK setting.

**Conclusions:** The CUAs for CHC were found to apply various HSUVs from different utility elicitation studies in the same health state. This variability of HSUVs has the potential to significantly affect ICER and ICER-based reimbursement decision. A rigorous selection of HSUVs in CUAs to inform healthcare resource allocation is suggested for future studies of CUAs and guideline development.

## Introduction

Hepatitis C is a slowly progressing infectious disease of the liver arising from the hepatitis C virus (HCV). The burden of disease of chronic hepatitis C (CHC) is substantial in Europe [1] with more than 14 million people living with chronic HCV infection [2]. The seroprevalence of HCV infection varies widely between European countries among the general population and high-risk populations including people who inject drugs (PWID) and men who have sex with men (MSM) [3]. The dramatic improvement in sustained virological response (SVR) rates, once-daily dosing, and short therapies with interferon-free direct acting antiviral therapies (IFN-free DAAs) lead to the speculation that CHC treatment could feasibly be scaled-up sufficiently and target groups could be broadened [4]. However, the high acquisition costs of DAAs limit the access for patients and influence the costs of healthcare resource utilisation in CHC [5-7]. There have been a growing number of pharmacoeconomic evaluations performed in CHC to inform healthcare resource allocation [8-18].

Health state utility values (HSUVs) are widely applied in pharmacoeconomic evaluations. HSUVs are used as weights in the calculation of the quality-adjusted life-years (QALYs), which incorporates both quantity and quality of life and is considered as an ideal measure of health outcomes in pharmacoeconomic evaluations [19]. Cost-utility analysis (CUA), a type of pharmacoeconomic evaluation which uses incremental cost per QALY gained (ICER) as a health outcome, is preferred in many countries requiring health technology assessment (HTA) for drug reimbursement [20], such as in the United Kingdom (UK) [21], France [22], Canada [23], Japan [24] and recently in China [25].

Although many CUAs were conducted for pharmacoeconomic evaluations in CHC [13], different HSUVs associated with the same health state were applied in the estimation of QALYs gained [9]. Xie et al. [26] summarised the current issues related to HSUVs used in CUAs into the following 3 domains: 1) identification (large variations in HSUVs between studies), 2) quality (limited methodological standards for critical appraisal), and 3) appropriate use (context-specific process of reimbursement decision making). The heterogeneity in applying HSUVs due to the different selection of utility elicitation studies with different elicitation methods, such as the visual analogue scale (VAS), the standard gamble (SG), time trade-off (TTO) and indirect methods by EQ-5D and SF-6D etc., and different responders, such as patient, physician and layperson. The impact of HSUVs selection on resulting ICER were addressed by Zhou et al. [27] using the case of schizophrenia. However, the situation of identification and selection of HSUVs in CUAs for CHC remains unclear and the impact on ICER has not been investigated. The objective of this study is to identify HSUVs used in the CUA models for CHC conducted in European countries and evaluate the impact of HSUVs selection on ICER.

## Methods

### Systematic review of pharmacoeconomic evaluation models

A recently published systematic review [8] of pharmacoeconomic evaluations for CHC was updated to identify CUAs conducted in European countries and reporting the HSUVs of CHC. The systematic search of the previous review for the period from 2012 to 2017 identified 2,917 studies and included 69 unique studies. The search results were updated for the period from 2017 to 2020.

#### Data source and data selection

Pharmacoeconomic evaluation models for CHC were retrieved from Medline and Embase for the period from 2017 to 2020. The search strategy is based on the previous review [8] and is presented in Additional file 1.1. The search terms covered the following domains: 1) population (patients with CHC or HCV infection), 2) intervention (pharmaceutical treatment), 3) evaluation technique (modelling) and 4) evaluation type (cost analysis or cost minimisation analysis or cost-effectiveness analysis or cost-utility analysis or cost-benefit analysis). The inclusion/exclusion criteria are presented in Additional file 1.2. Studies were included if 1) they were conducted in adult patients (aged 18 years) with CHC; 2) they evaluated all pharmaceutical treatment strategies of CHC including DAAs and interferon; 3) they were model-based economic evaluations; 4) published after 2017 to the present; 5) conducted in European countries. The identified models were pooled with the models identified from the previous review [8] for a further screening to include the CUAs conducted in European countries and reporting the HSUVs of CHC. References of the identified models were screened and included when applicable.

#### Data extraction and data synthesis

The extracted information from the included CUAs covered two domains: 1) study design including country, population, objective, perspective, cost year, model design, time horizon, uncertainty analysis, subgroup analysis, intervention and comparator, and outcome; 2) health states and HSUVs including health states and their definitions, utility values for each health state, associated source, and an interpretation of HSUVs from the source into the CUA model. The following information was summarised: number of CUAs considering the health state, number of HSUVs per health state, number of utility elicitation studies used as the source of HSUVs, and the utility values (most common value and range of values) for each health state of CHC. The difference in HSUVs of the same health state in different CUAs and the difference between HSUVs of one health state and of the interlink health state in the same CUAs were calculated. Utility values for all health states of CHC within each CUA model were considered as one set of HSUVs.

### Impact of HSUVs identification on ICER

#### Identification of utility elicitation study

The process of investigating the impact of HSUVs selection on cost-effectiveness results was reported by Zhou et al. [27]. The reported sources of HSUVs in included CUAs were screened to identify the utility elicitation studies which generated these HSUVs. If an associated source was another CUA, the references of this CUA would be screened. The following information was extracted from utility elicitation studies: country, population or responder, utility elicitation method and health states of CHC and their definitions when applicable.

#### Quality assessment of HSUVs used in CUAs

A framework to evaluate the identification and selection of HSUVs in CUAs was developed based on Xie et al. [26] and Nerich et al. [28]. Each included a CUA model and was evaluated for the following 3 domains covering 7 items: 1) identification of the source of HSUVs (selection based on systematic review and sensitivity analysis or reasonable justification); 2) methodological quality (HSUVs elicitation method and responder); 3) appropriate selection of HSUVs from the source (population comparability, health states comparability and discussion). Points of 0-2 were awarded in each item and indicated whether; no description (0), partial description (1), or complete description (2) in each item, was provided by the authors. The average point of all HSUVs in one CUA was calculated as the final point of the CUA in each item. More details of the quality assessment framework are presented in Additional file 1.3.

#### Evaluation of impact of HSUVs identification on ICER

A core Markov model from Shepherd et al. [29-31] over lifetime with one-year cycle length was used to investigate the impact of HSUVs selection on the cost-effectiveness result, which is reported as ICER in the CUA for CHC. This Markov model was chosen because it was widely used with many adaptations [32-42]and easy to replicate based on its simplicity of approach and completeness of reporting in the publication. The model structure is presented in Figure 1: 1) 6 core health states including chronic HCV (Metavir F0-F3), compensated cirrhosis (Metavir F4), decompensated cirrhosis, hepatocellular carcinoma, liver transplant in the first year and liver transplant in subsequent years; 2) 2 recovered states including SVR from chronic HCV infection without cirrhosis and SVR from compensated cirrhosis; 3) death (absorbing health states for liver and non-liver disease). The baseline characteristics, transition probability, clinical efficacy and costs were obtained from Johnson et al. [43], a CUA of ombitasvir/paritaprevir/ritonavir and dasabuvir ± ribavirin (ODR) versus pegylated interferon + ribavirin (PR) conducted in the UK, and are presented in Additional file 1.4. The HSUVs for 6 core health states were selected from the identified sets of HSUVs. The utility values for recovered states were not considered in the present analysis since they were rather collected from trials than elicited using utility weights to synthesise the multi-dimension impact of treatment [44]. To fit the HSUVs sets identified in all CUAs included, the following modifications were made: 1) when combined health states, such as a combined cirrhosis state of decompensated cirrhosis and compensated cirrhosis, and a combined liver transplant state of the first year and subsequent years, were used, each individual health state was assumed to have the same HSUV as the combined health state; 2) when multiple HSUVs were reported for one health state, the average value was used.

**Figure 1.**
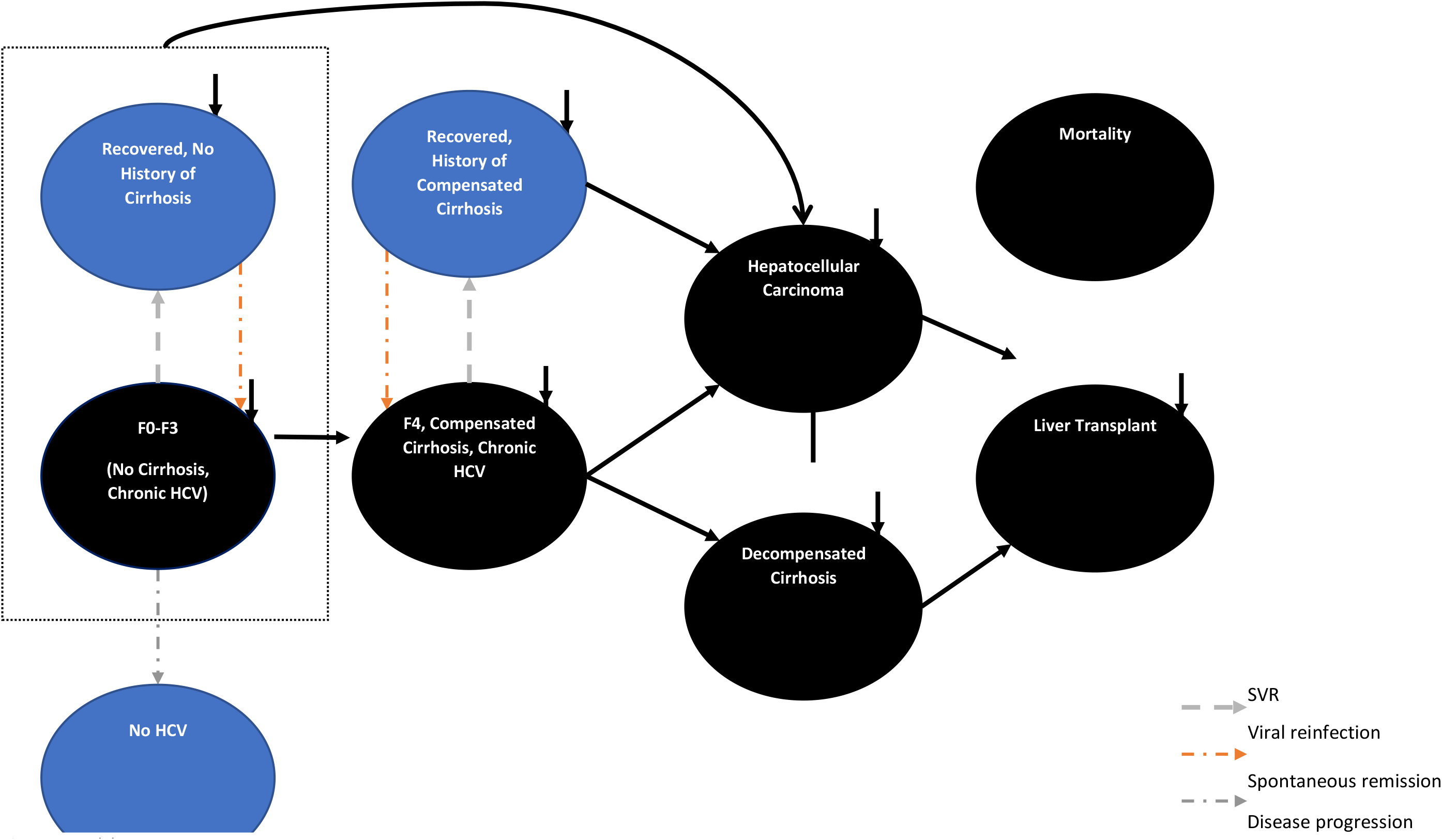
Model structure

## Results

### Systematic review of pharmacoeconomic evaluation models

After the title/abstract screening of 923 publications, the present review identified an additional 39 pharmacoeconomic evaluation models (Figure 2) conducted in European countries which were not included in the 15 models identified in the previous review [8]. Out of 54 studies, 26 CUAs [18, 43, 45-68] reporting detailed HSUVs of CHC were included in the analysis of impact of HSUVs selection on ICER and the other 28 were excluded because they were not CUAs or did not report detailed HSUVs. The characteristics of all CUAs included (n=26) in the study design are presented in Table 1. The total CUAs included were conducted in 7 European countries (mostly in the UK, n=8). Most CUAs used the cohort-level Markov model (n=13), took the perspective of a public healthcare payer (n=22), adopted lifetime horizon (n=20) and reported ICER (n=24).

**Table 1.** Summary of characteristics in study design of all CUA models included

| Comparison area | Description | n |
| --- | --- | --- |
| Type of model | Cohort-level Markov model | 13 |
|  | Dynamic transmission model | 6 |
|  | Multicohort Markov model | 4 |
|  | Semi-Markov | 1 |
|  | Decision tree and Markov model | 1 |
|  | Econometric model | 1 |
| Country | UK | 8 |
|  | Italy | 8 |
|  | Spain | 3 |
|  | France | 2 |
|  | Germany | 2 |
|  | Netherlands | 2 |
|  | Norway | 1 |
| Perspective | Payer | 22 |
|  | Healthcare provider | 1 |
|  | Societal | 1 |
|  | Payer and patient | 1 |
|  | NR | 1 |
| Time horizon | Lifetime | 20 |
|  | 50 years | 3 |
|  | 30 years | 1 |
|  | 15 years | 1 |
|  | NR | 1 |
| Patient population | General patient with chronic hepatitis C | 21 |
|  | PWID with chronic hepatitis C | 3 |
|  | HIV-infected MSM with chronic hepatitis C | 1 |
|  | HIV-infected patient with chronic hepatitis C | 1 |
| Type of comparison | Pharmacotherapy | 23 |
|  | Treatment timing/priority | 8 |
|  | Treatment uptake rate level | 1 |
|  | HCV prevalence level | 1 |
|  | SVR level | 1 |
| Outcomes | Time spent in status |  |
|  | • QALYs gains | 26 |
|  | • LYs gains | 7 |
|  | Number/percentage of patients with events* | 7 |
|  | Mortality outcomes | 1 |
|  | Cost outcomes | 26 |
|  | Economic outcomes |  |
|  | • incremental cost per QALY gained | 24 |
|  | • incremental net monetary benefit | 5 |
|  | • incremental cost per life-year gained | 1 |
|  | • maximum acceptable financial expenditure | 1 |

\*Events represented advanced liver complications including compensated cirrhosis, decompensated cirrhosis, hepatocellular carcinoma and liver transplant or dying from liver-related causes. CUAs: cost
utility analyses; LYs: life-years; NR: not reported; QALYs: quality-adjusted life-years; SVR: sustained virological response; UK: United Kingdom

**Figure 2.**
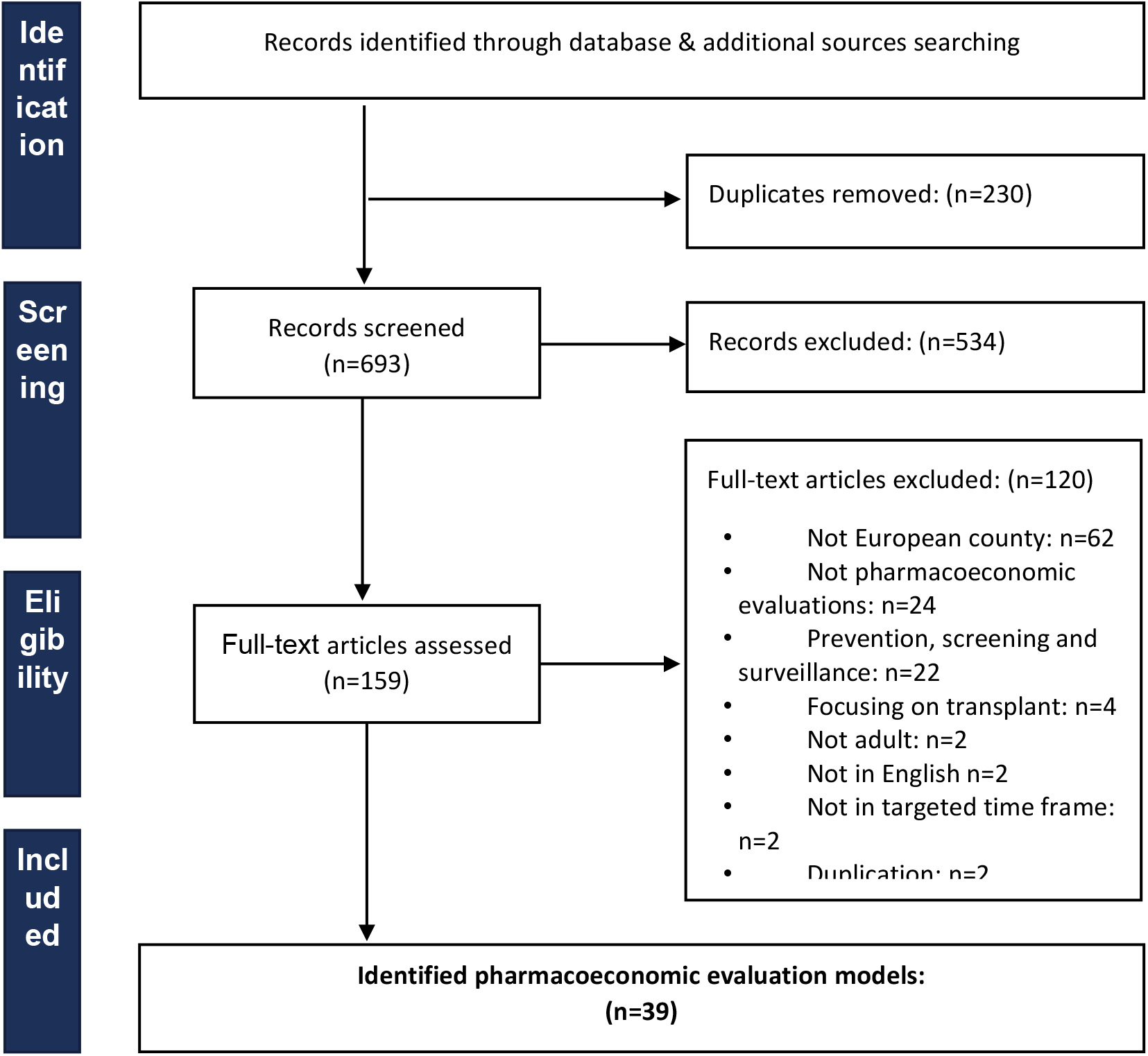
PRISMA diagram of selection process of pharmacoeconomic evaluations of chronic hepatitis C

### Characteristics of health states and HSUVs used in CUAs

The characteristics of all CUAs included (n=26) in health states and HSUVs are presented in Table 2. Expanded health states of early chronic hepatitis and combined health states of advanced liver complications to the 6-health-state core model [29-31] were identified in most CUAs: 1) classifications of health states of early chronic hepatitis: F0-F3/chronic HCV infection, F4/compensated cirrhosis (n=6); F0-F1/mild, F2-F3/moderate, F4/compensated cirrhosis (n=5); F0, F1, F2, F3 (n=5); F0-F1, F2, F3-F4 (n=4); F0, F1-F2, F3, F4 (n=4); F0-F1, F2, F3, F4 (n=1); and F0-F2, F3-F4 (n=1); 2) classifications of health states of advanced liver complications: decompensated cirrhosis, hepatocellular carcinoma, liver transplant in the first year, liver transplant in subsequent years (n=17); decompensated cirrhosis, hepatocellular carcinoma, liver transplant(n=6); cirrhosis, hepatocellular carcinoma, liver transplant (n=1); and decompensated cirrhosis, liver transplant (n=1). Overall there were 10 health states of early chronic hepatitis and 5 health states of advanced liver complications. The most numbers of HSUVs were reported in the health state of decompensated cirrhosis (n=24, 0.380-0.848), hepatocellular carcinoma (n=24, 0.380-0.867), liver transplant in the first year (n=17, 0.450-0.852), liver transplant in subsequent years (n=17, 0.450-0.910), and F4/cirrhosis (n=14, 0.550-0.877). The difference in HSUVs of the same health states in different CUAs ranged from 0.021 (liver transplant) to 0.468 (decompensated cirrhosis). The difference between HSUVs of one health state and of the interlink health state was calculated between health state of F0-F1/mild and F2-F3/moderate (n=11, 0.040-0.110), F2-F3/moderate and F4/compensated cirrhosis (n=18, 0.027-0.130), compensated cirrhosis and decompensated cirrhosis (n=22, 0.020-0.100), decompensated cirrhosis and hepatocellular carcinoma (n=24, 0.000-0.200), hepatocellular carcinoma and liver transplant in the first year (n=17, −0.329-0.170) and liver transplant in the first year and in subsequent years (n=17, −0.340-0.000).

**Table 2.**
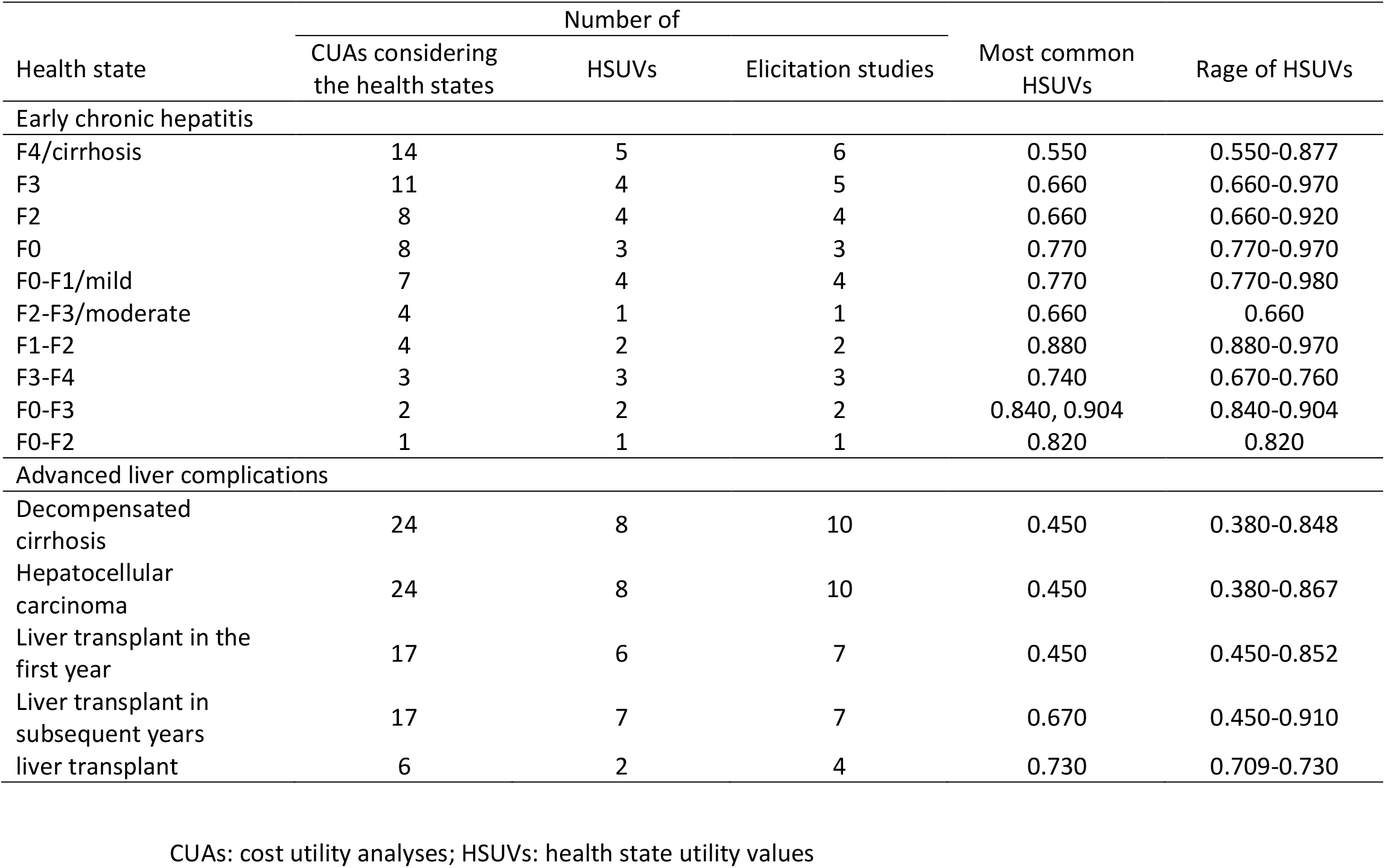
HSUVs of chronic hepatitis C in all CUA models included

| Health state | Number of |  |  | Most common HSUVs | Rage of HSUVs |
| --- | --- | --- | --- | --- | --- |
|  | CUAs considering the health states | HSUVs | Elicitation studies |  |  |
| Early chronic hepatitis |  |  |  |  |  |
| F4/cirrhosis | 14 | 5 | 6 | 0.550 | 0.550-0.877 |
| F3 | 11 | 4 | 5 | 0.660 | 0.660-0.970 |
| F2 | 8 | 4 | 4 | 0.660 | 0.660-0.920 |
| F0 | 8 | 3 | 3 | 0.770 | 0.770-0.970 |
| F0-F1/mild | 7 | 4 | 4 | 0.770 | 0.770-0.980 |
| F2-F3/moderate | 4 | 1 | 1 | 0.660 | 0.660 |
| F1-F2 | 4 | 2 | 2 | 0.880 | 0.880-0.970 |
| F3-F4 | 3 | 3 | 3 | 0.740 | 0.670-0.760 |
| F0-F3 | 2 | 2 | 2 | 0.840, 0.904 | 0.840-0.904 |
| F0-F2 | 1 | 1 | 1 | 0.820 | 0.820 |
| Advanced liver complications |  |  |  |  |  |
| Decompensated cirrhosis | 24 | 8 | 10 | 0.450 | 0.380-0.848 |
| Hepatocellular carcinoma | 24 | 8 | 10 | 0.450 | 0.380-0.867 |
| Liver transplant in the first year | 17 | 6 | 7 | 0.450 | 0.450-0.852 |
| Liver transplant in subsequent years | 17 | 7 | 7 | 0.670 | 0.450-0.910 |
| liver transplant | 6 | 2 | 4 | 0.730 | 0.709-0.730 |

CUAs: cost utility analyses; HSUVs: health state utility values

### Impact of HSUVs selection on ICER

#### Quality assessment of HSUVs used in CUAs

In total, 17 utility elicitation studies [9, 40, 57, 69-82] and 7 sets of HSUVs of CHC were used in all CUAs included. The characteristics of all utility elicitation studies included are presented in Table 3. Wright et al. [69] was the most referred study (n=11) for health states of mild, moderate, compensated cirrhosis, decompensated cirrhosis, hepatocellular carcinoma, liver transplant in the first year and subsequent years with high comparability of population and between health states in utility elicitation studies and in CUAs. Most CUAs (n=22) derived the HSUVs from a single utility elicitation study, and the others (n=6) derived them from multiple sources. The results of quality assessment of HSUVs used in CUAs for CHC are presented in Table 4. Most CUAs (n=25) identified HSUVs from literatures and reported the sources and values and 2 of them identified HSUVs through a systematic review; only one CUA used assumed HSUVs. Most CUAs (n=21) conducted sensitivity analysis or scenario analysis of alternative HSUVs and only 2 of them did not report the range or source of alternative HSUVs. Most CUAs (n=20) did not describe the elicitation method and responder (n=23); the others provided an incomplete description without an explanation of choice of elicitation method and responder. Some CUAs (n=11) used the utility elicitation studies with the same population and country as in CUAs; the others used a different population, such as a population with a different diagnosis, severity level and risk behaviour, and different country. Half CUAs (n=13) used health states which were different from that applied in elicitation studies without an explanation and 5 CUAs used different health states but with an explanation on derivation method. Most CUAs (n=17) conducted an incomplete discussion on the limitations of the HSUVs source identification, the elicitation method and HSUVs selection; the others provided no discussion on HSUVs at all.

**Table 3.** Characteristics of the utility elicitation studies referred in all CUA models included

| Author Year | Country | Elicitation method | Responder | Health states |
| --- | --- | --- | --- | --- |
| Wright 2006 [69] | UK | EQ-5D | patient | mild, moderate, cirrhosis, HCC, treatment for mild, treatment for moderate, decompensated cirrhosis, post-liver transplantation |
| Kind 2008 [70] | UK | EQ-5D | resident | uninfected individual |
| Hzode 2014 [71] | international multicentre | NR | NR | NR |
| Sulkowski 2014 [72] | US | NR | NR | NR |
| McDonald 2013 [73] | UK | EQ-5D | patient | chronically infected and aware of infected status, chronically infected but unaware, not chronically infected |
| Townsend 2011 [9] | international multicentre | NR | NR | mild, moderate, F0-F3, compensated cirrhosis, decompensated cirrhosis, hepatocellular carcinoma, transplant in the first year, transplant in subsequent years |
| McLernon 2008 [74] | international multicentre | EQ-5D | patient | moderate, compensated cirrhosis, decompensated cirrhosis, and post-liver transplant |
| Sieber 2003 [75] | Germany | visual analogue scales | patient | mild, moderate, decompensated cirrhosis, hepatocellular carcinoma, liver transplantation |
| Chong 2003 [76] | Canada | standard gamble | patient | no cirrhosis, mild to moderate chronic HCV infection, compensated cirrhosis, decompensated cirrhosis, hepatocellular carcinoma, transplant |
| Cossais 2015 [77] | France | EQ-5D | patient | F0, F1, F2, F3, F4 |
| Pol 2015 [82] | France, the UK and Germany | EQ-5D | patient | F0, F1, F2, F3, F4, decompensated cirrhosis, hepatocellular carcinoma, liver transplant in the first year, liver transplant in subsequent years |
| Kondil 2017 [57] | Italy | NR | NR | F0, F1-F2, F3, F4, decompensated cirrhosis, hepatocellular carcinoma, liver transplant in the first year, liver transplant in subsequent years |
| Sullivan 2004 [40] | US | HUI 3 | patient | chronic hepatitis C, compensated cirrhosis, decompensated cirrhosis, hepatocellular carcinoma and liver transplantation |
| Tammy 2002 [78] | international multicentre | time trade-off | patient | asymptomatic HIV |
| McGrealBellone 2012 [79] | Ireland | EQ-5D | patient | HIV/HCV coinfectd patient with cirrhosis, without cirrhosis |
| Cortesi 2013 [80] | Italy | EQ-5D | patient | NR |
| Sullivan 2006 [81] | US | EQ-5D | patient | NR |

\*EQ-5D scores were derived from SF-6D using mapping algorithm. HIV: human immunodeficiency viruses; NR: not reported; UK: United Kingdom; US: United States

**Table 4.**
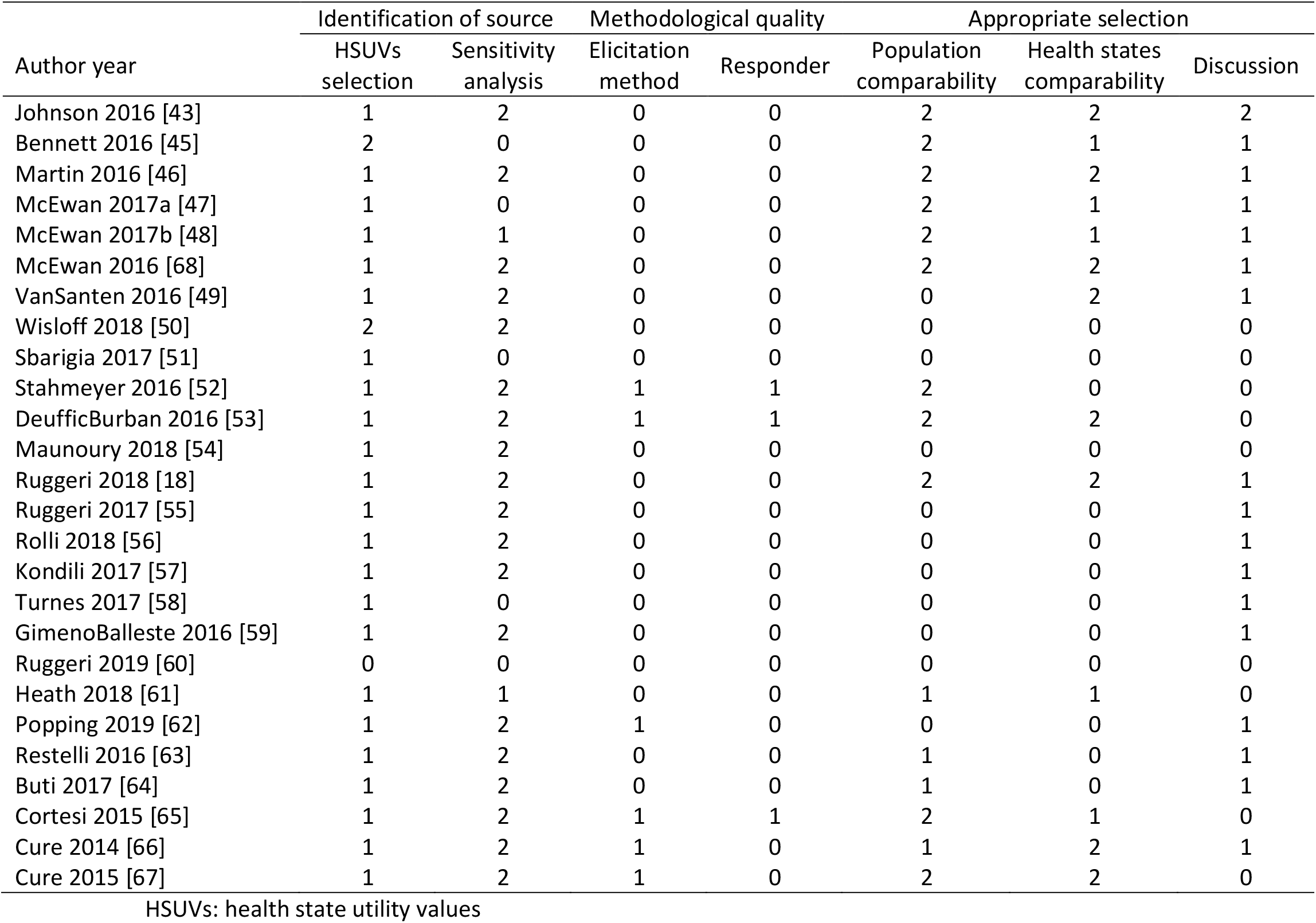
Quality assessment of HSUVs used in all CUA models included

| Author year | Identification of source |  | Methodological quality |  | Appropriate selection |  |  |
| --- | --- | --- | --- | --- | --- | --- | --- |
|  | HSUVs selection | Sensitivity analysis | Elicitation method | Responder | Population comparability | Health states comparability | Discussion |
| Johnson 2016 [43] | 1 | 2 | 0 | 0 | 2 | 2 | 2 |
| Bennett 2016 [45] | 2 | 0 | 0 | 0 | 2 | 1 | 1 |
| Martin 2016 [46] | 1 | 2 | 0 | 0 | 2 | 2 | 1 |
| McEwan 2017a [47] | 1 | 0 | 0 | 0 | 2 | 1 | 1 |
| McEwan 2017b [48] | 1 | 1 | 0 | 0 | 2 | 1 | 1 |
| McEwan 2016 [68] | 1 | 2 | 0 | 0 | 2 | 2 | 1 |
| VanSanten 2016 [49] | 1 | 2 | 0 | 0 | 0 | 2 | 1 |
| Wisloff 2018 [50] | 2 | 2 | 0 | 0 | 0 | 0 | 0 |
| Sbarigia 2017 [51] | 1 | 0 | 0 | 0 | 0 | 0 | 0 |
| Stahmeyer 2016 [52] | 1 | 2 | 1 | 1 | 2 | 0 | 0 |
| DeufficBurban 2016 [53] | 1 | 2 | 1 | 1 | 2 | 2 | 0 |
| Maunoury 2018 [54] | 1 | 2 | 0 | 0 | 0 | 0 | 0 |
| Ruggeri 2018 [18] | 1 | 2 | 0 | 0 | 2 | 2 | 1 |
| Ruggeri 2017 [55] | 1 | 2 | 0 | 0 | 0 | 0 | 1 |
| Rolli 2018 [56] | 1 | 2 | 0 | 0 | 0 | 0 | 1 |
| Kondili 2017 [57] | 1 | 2 | 0 | 0 | 0 | 0 | 1 |
| Turnes 2017 [58] | 1 | 0 | 0 | 0 | 0 | 0 | 1 |
| GimenoBalleste 2016 [59] | 1 | 2 | 0 | 0 | 0 | 0 | 1 |
| Ruggeri 2019 [60] | 0 | 0 | 0 | 0 | 0 | 0 | 0 |
| Heath 2018 [61] | 1 | 1 | 0 | 0 | 1 | 1 | 0 |
| Popping 2019 [62] | 1 | 2 | 1 | 0 | 0 | 0 | 1 |
| Restelli 2016 [63] | 1 | 2 | 0 | 0 | 1 | 0 | 1 |
| Buti 2017 [64] | 1 | 2 | 0 | 0 | 1 | 0 | 1 |
| Cortesi 2015 [65] | 1 | 2 | 1 | 1 | 2 | 1 | 0 |
| Cure 2014 [66] | 1 | 2 | 1 | 0 | 1 | 2 | 1 |
| Cure 2015 [67] | 1 | 2 | 1 | 0 | 2 | 2 | 0 |

HSUVs: health state utility values

#### Evaluation of impact of HSUVs selection on ICER

After the modification of HSUVs based on the predefined rules to fit the model analysis, 7 sets of HSUVs of CHC in the re-constructed model [29-31] were generated. The selection of different HSUVs sets in the model changed the results of incremental analysis from being cost-effective to not cost-effective, and to dominated in the UK setting (Table 5). an incremental QALY increasing from 0.028 to 1.635 and an ICER raging from £3,483 to £200,940 per QALY gained in 3 HSUVs sets and decreasing from −1.270 to −0.607 in the other 4 HSUVs sets.

**Table 5.** Impact of different HSUV sets of chronic hepatitis C

| HSUVs set | HSUV |  |  |  |  |  | Incremental QALY | ICER (£) |
| --- | --- | --- | --- | --- | --- | --- | --- | --- |
|  | Chronic HCV | Compensated cirrhosis | Decompensated cirrhosis | Hepatocellular carcinoma | Liver transplant (first year) | Liver transplant (subsequent) |  |  |
| Wright 2006 [69] | 0.715* | 0.550 | 0.450 | 0.450 | 0.450 | 0.67 | 1.635 | 3,483 |
| Townsend 2011 [9] | 0.747 | 0.733# | 0.733# | 0.380 | 0.709# | 0.709# | 0.673 | 8,467 |
| Cossais 2015 [77] | 0.820 | 0.760# | 0.760# | 0.600 | 0.550 | 0.820 | 0.028 | 200,940 |
| Sieber 2003 [75] | 0.935* | 0.890 | 0.810 | 0.810 | 0.860 | 0.860 | -1.254 | Dominated |
| Kondil 2017 [57] | 0.970 | 0.830 | 0.730 | 0.530 | 0.730# | 0.730# | -1.270 | Dominated |
| Sullivan 2004 [40] | 0.880 | 0.830 | 0.730 | 0.530 | 0.730# | 0.730# | -0.607 | Dominated |
| Cortesi 2013 [80] | 0.904 | 0.877 | 0.848 | 0.867 | 0.852 | 0.910 | -1.011 | Dominated |

\*Calculated as the average value of mild and moderate chronic HCV based on modification rules.
#Assumed to equal to the HSUV of cirrhosis and liver transplant based on modification rules. HCV: hepatitis C virus; HSUVs: health state utility values; ICER: incremental cost per quality-adjusted life-year (QALY) gained; QALYs: quality-adjusted life-years

## Discussion

This study reviewed the HSUVs used in the CUA models for pharmacoeconomic evaluations of CHC and investigated the impact of HSUVs selection on resulting ICER. The CUAs for CHC were found to apply various HSUVs from different utility elicitation studies. This variability has the potential to significantly affect ICER. The findings of this study suggest that the HSUVs sources on which pharmacoeconomic evaluations are based on may not adequately reflect the patient population and may not be sufficient for the needs of current research questions, as long as there is no standard to identify and apply the utility elicitation studies and HSUVs to the CUAs.

In the CUAs included by this systematic review, 6-health-state core model of CHC were widely applied: F0-F3/chronic HCV infection, F4/compensated cirrhosis, decompensated cirrhosis, hepatocellular carcinoma, liver transplant in the first year and liver transplant in subsequent years. Two common variations to core health states were identified: 1) expansion of chronic HCV infection health state to reflect varying levels of disease severity at model entry, either using broad categories such as “mild” and “moderate” or using a Metavir scoring system based on histological status of fibrosis; 2) combination of health states of decompensated cirrhosis and compensated cirrhosis, and liver transplant in the first year and in subsequent years into health states of cirrhosis and liver transplant. The first variation of health state of chronic HCV infection is consistent with a previous review [9] of structural frameworks of CUAs for CHC. Although some CUAs applied core health states with adaptations and variations, the underlying HSUVS are the same and derived from the same utility elicitation study. The value of the expansion of the early chronic HCV infection is to assess the cost-effectiveness of early treatment with antivirals at a time of higher SVR [9]. However, using the same HSUVs and utility elicitation studies and applying optimistic assumption without justification render the expansion of core health states worthless.

There was a high level of variation in HSUVs of the same health state in CHC applied in different CUAs. However, the relatively small difference between HSUVs of one health state and the interlink health state in the same CUAs was identified. No systematic pattern was found regarding how HSUVs have been selected from different utility elicitation studies and how the variation is distributed across the study. The utility elicitation studies differed in countries, elicitation methods, responders and classifications of health states of CHC. The HSUVs used in the CUAs frequently do not directly reflect the country-specific perspective of the research question and are derived from the different health states with different interpretations in the utility elicitation study and the CUA. Of all utility elicitation studies, Wright et al. [69] was most referred, where the HSUVs for patients at each predefined health state were collected as part of the multicentre, randomised, controlled, non-blinded trial in the UK National Health System setting and estimated using EQ-5D questionnaire. The descriptions of each patient’s health status were then translated into HSUV using a reference set of preference weights derived from a representative sample of the UK general population using the TTO technique. Compared with estimates from expert opinions and elicitation studies conducted in the other countries and with undefined health states, this study elicited responses from patients to estimate the HSUVs for the main health states included in the CUA model.

In the absence of utility elicitation studies conducted among high-risk populations such as PWID and MSM, most CUAs [45, 46] assumed that HSUVs were comparable between PWID/MSM and non-PWID/non-MSM which was not consistent with the reported result that there was a lower quality of life in terms of utility among high-risk populations [83]. One CUA among PWID [49] applied HSUVs in the general population multiplied by 0.85 to account for drug dependency. The limitation and validity of HSUVs should be fully discussed to interpret the results in CUAs given that the ICER achieved is significantly influenced by the HSUVs.

It is noteworthy that ICER varies considerably with different set of HSVUs when applying the exact same model, assumptions and input for the other parameters. The level of difference in ICER could significantly influence ICER-based reimbursement decisions. The variability in HSUVs incorporated in this CUA can change the resulting ICER of DAA versus PR from being cost-effective at the common UK threshold value of £20,000 per QALY gained to dominated, when all other considerations remain equal. These results are in line with results from Townsend et al. [9].

Given the results, a quality assessment of the identification and selection of utility elicitation studies and HSUVs in CUAs for CHC was applied. The value of the quality assessment is to serve as a guide of rigorous conduct and proper use of HSUVs for future CUAs and generate disease-specific evidence for the currently developed systematic approach to appropriate use of HSUVs in CUAs [26].

This study has several limitations. First, the systematic review of CUAs focused on analyses conducted in European countries. It is possible that a difference between studies in country perspective may affect results. However, it is not necessary to control for possible country-specific variation because the underlying HSUVs sources were rarely directly relevant to the country perspective and commonly taken from pooled international studies. Next, modifications were made when the health states in the utility elicitation study did not completely match that used in the re-constructed model. However, these modifications still captured the major difference in HSUVs between health states when compared with HSUVs without modification. Last, a simplistic quality assessment of HSUVs used in CUAs was conducted. In the absence of rigorous standard for the identification and selection of utility elicitation studies and HSUVs, these results can provide guidance for future CUAs and the development of guidelines in pharmacoeconomic evaluations.

## Conclusion

The CUAs for CHC were found to apply various HSUVs from different utility elicitation studies on the same health state. This variability of HSUVs has the potential to significantly affect ICER and ICER-based reimbursement decision. A rigorous identification and selection of HSUVs in CUAs to inform healthcare resource allocation is suggested for future studies of CUAs and guideline development.

## Data Availability

The data relating to the systematic review are all included in the article and/or its supplementary materials. The model used in the study can be made available to researchers upon request.

## Acknowledgements

Not applicable.

## Permissions

Not applicable.

## Declarations

### Ethics approval

Not applicable.

### Consent to participate

Not applicable.

### Consent for publication

Not applicable.

### Availability of data and material

Not applicable.

### Code availability

Not applicable.

### Contributors

All authors contributed to study conception and design. Ru Han contributed to the literature search, acquisition of data, analysis and interpretation of data and first draft of the manuscript. Clément François and Junwen ZHOU contributed to critical revision of the manuscript for important intellectual content. Clément François and Mondher Toumi contributed to supervision of the study. Marcus Bashford contributed to writing or technical editing of the manuscript. All authors read and approved the final manuscript.

